# What factors increase the risk of complications in SARS-CoV-2 positive patients? A cohort study in a nationwide Israeli health organization

**DOI:** 10.1101/2020.05.07.20091652

**Authors:** Chen Yanover, Barak Mizrahi, Nir Kalkstein, Karni Marcus, Pinchas Akiva, Yael Barer, Varda Shalev, Gabriel Chodick

**Affiliations:** KI Research Institute, Kfar Malal, Israel; Maccabi Institute for Research and Innovation, Tel Aviv, Israel; Sackler Faculty of Medicine, Tel-Aviv University, Tel-Aviv, Israel

## Abstract

Reliably identifying patients at increased risk for COVID-19 complications could guide clinical decisions, public health policies, and preparedness efforts. To date, the most globally accepted definitions of at-risk patients rely, primarily, on epidemiological characterization of hospitalized COVID-19 patients. However, such characterization overlooks, and fails to correct for, the prevalence of existing conditions in the wider SARS-CoV-2 positive population. Here, we analyze the complete medical records of all SARS-CoV-2 infected individuals (N=4,353) in a large Israeli health organization (representing a population of 2.3 million people), of whom 173 experienced moderate or severe symptoms of COVID-19, to identify the conditions that increase the risk of disease complications, in various age and sex strata. Our analysis suggests that cardiovascular and kidney diseases, obesity, and hypertension are significant risk factors for COVID-19 complications, as previously reported. Interestingly, it also indicates that depression (e.g., odds ratio, OR, for males 65 years or older: 2.94, 95% confidence intervals [1.55, 5.58]; P-value = 0.014) as well cognitive and neurological disorder (e.g., OR for individuals ≥ 65 year old: 2.65 [1.69, 4.17]; P-value < 0.001) are significant risk factors; and that smoking and background of respiratory diseases do not significantly increase the risk of complications. Adjusting existing risk definitions following these observations may improve their accuracy and impact the global pandemic containment efforts.

## Introduction

As of May 7^th^, 2020, close to four million people worldwide contracted severe acute respiratory syndrome coronavirus 2 (SARS-CoV-2) and more than 265,000 people died of corona virus disease 2019 (COVID-19) complications. This pandemic poses grave challenges to patients, healthcare providers, and policy makers. Many of these challenges may be better addressed with timely stratification of patients to risk groups, based on their past and current medical characteristics. For example, reliably identifying patients at increased (or decreased) risk could guide clinical decisions (e.g., hospitalization vs home care), public health policies (e.g., risk-based quarantine), and preparedness efforts (e.g., expected medical equipment required).

Various algorithms for identifying patients at risk for COVID-19 (severe) complications have been proposed. The Centers for Disease Control and Prevention (CDC) identified individuals 65 years and older, living at nursing home or long-term care facility, or suffering from underlying medical conditions, particularly if not well controlled, as being at high risk for severe illness from COVID-19 [1]. Similarly, the European Centre for Disease Prevention and Control (ECDC) lists age above 70 years and some underlying conditions as risk factors for critical illness [2]. The United Kingdom National Health Service (NHS) included solid organ transplant recipients, patient with specific cancers or severe respiratory conditions, pregnant women with significant heart disease, and those with increased risk of infection (e.g., due to immunosuppression therapies) in the highest clinical COVID-19 risk group [3]. In April 2020 approximately 1.3 million people in this group were asked to "shield" by staying at home for a period of at least 12 weeks. In addition, patients over 70 years and those suffering from some underlying health conditions (e.g., chronic respiratory diseases, BMI ≥ 40, and pregnant women) were considered in a wider vulnerable group (also referred to as the "flu group"). Finally, a more quantitative risk model (derived from [4]) has been adopted by the Israeli Ministry of Health (MoH), assigning a point for each underlying condition from a predefined list, then considering age group and point count to identify high risk patients.

Initially, these algorithms have been derived from a quickly growing number of epidemiological characterization studies (e.g., [5,6]), which report the prevalence of various conditions in a population of interest, typically severe COVID-19 patients. These studies provide timely and important information; however, identifying risk factors calls for a comparative analysis, contrasting the prevalence of conditions in case and control populations. To date, only a handful of studies implemented such an approach, using, for example, the general population [7] or confirmed (and symptomatic) COVID-19 patient cohort [8]. Similar to these efforts, we analyze here the medical records of all SARS-CoV-2 positive patients in a nationwide health organization (covering 2.3 million individuals); compare the prevalence of existing conditions in complicated and non-complicated cohorts; and identify those conditions associated with COVID-19 complications in various age and sex strata. Our analysis highlights stratum-specific risk factors and may allow better identification of patients at risk, in different subpopulations.

## Methods

### Maccabi COVID-19 data

Maccabi Health Services (MHS) is a nationwide health plan (payer-provider), representing a quarter of the Israeli population. The MHS database contains longitudinal data on a stable population of over 2.3 million people since 1993 (with annual attrition rate lower than 1%). Data are automatically collected and include comprehensive laboratory data from a single central lab, full pharmacy prescription and purchase data, and extensive demographic information on each patient.

### Studied cohorts

SARS-CoV-2 polymerase chain reaction testing in Israel uses both nasopharyngeal and saliva samples. Individuals with positive testing result (until April 22, 2020) are included in the *SARS-CoV-2 positive cohort*. Positive patients whose disease status, as updated by Israeli hospitals, deteriorated to moderate or severe (at any point in time), admitted to the intensive care unit, or died constitute the *complicated COVID-19 cohort;* the remaining SARS-CoV-2 positive patients (including asymptomatic, mild COVID-19 patients or those with unknown status) constitute the *non-complicated COVID-19 cohort*.

### Existing conditions

Beside age and sex, we considered a set of existing conditions, comprising those included in the CDC, NHS, and Israeli MoH at-risk definitions, as well as a set of conditions showing significant association with flu and flu-like complications.

To identify each individual’s existing conditions, we used, where available, registries created and maintained by MHS. These registries are based on validated inclusion and exclusion criteria (considering coded diagnoses, treatment, labs, and imaging, as applicable). The registries are continuously and retrospectively (since 1998) updated based on each patient’s central medical record. Patients may be excluded from a registry when deemed misclassified by their primary physician. Linkage across registries and with other sources of information is performed via a unique national identification number. MHS registries used are: Cardiovascular diseases (including ischemic heart disease, congestive heart failure, peripheral vascular disease, cerebrovascular disease, and other cardiovascular diseases) [9], diabetes [10,11], hypertension [12], osteoporosis [13], chronic kidney disease [14], cognitive disorders, mental illness [15], cancer, immunosuppression, weight disorders (obesity, overweight and underweight), smoking, and nursing home. For other conditions, we relied on previously grouped lists of diagnosis codes (Read codes or International Classification of Diseases, ICD, codes, ninth revision) [16-18]: Deficiency anemia, Fluid and electrolyte disorders, chronic obstructive pulmonary disease (COPD), chronic pulmonary disease, neurological disorders, end stage renal disease, rheumatoid arthritis, paralysis, hip fracture, lymphoma, aspiration pneumonia, pleural effusion, respiratory failure, and alcohol consumption.

### Statistical analysis

We extracted the prevalence of the studied conditions (excluding ones with less than 20 occurrences) in the non-complicated and complicated COVID-19 cohorts and measured the association between each condition and disease complications by computing the corresponding odds ratio and its estimated statistical significance (using Fisher’s exact test). We conducted the analysis separately in three age groups: 18-50 years, 50-65 years, and 65 years and older; as well as four (age, sex) strata: male or female, younger or older than 65 years. Finally, to account for multiple testing, we controlled for the false discovery rate using Benjamini and Hochberg’s method [19]. All analyses were performed using version 4.0.0 of the R programming language (R Project for Statistical Computing; R Foundation).

## Results

Maccabi Health Services (MHS) SARS-CoV-2 positive cohort included 4,353 individuals of whom 173 deteriorated to moderate (N=87, 50%) or severe condition (N=45, 26%), admitted to intensive care unit (N=66, 38%, partly overlapping with other conditions), or died (N=21, 12%), and make up the complicated COVID-19 cohort. Overall, patients in the complicated COVID-19 cohort are older, suffer from more comorbidities, and are more predominantly male (Table 1). Moreover, the prevalence of COVID-19 complications increases with age, and more steeply for men than for women (Figure 1.); and the risk of COVID-19 complications in men under 70 years is significantly higher than in women (Table 2).

**Table 1.** Characteristics of the SARS-CoV-2 positive, complicated, and non-complicated COVID-19 patient cohorts.

|  | SARS-Cov-2 positive | Complicated COVID-19 | Non-complicated COVID-19 |
| --- | --- | --- | --- |
| **Demographic information** |  |  |  |
| N | 4353 | 173 | 4180 |
| Age median [inter quantile range, IQR] | 35 [22-54] | 70 [58-80] | 34 [22-52] |
| <18y | 647 (15%) | 0 (0%) | 647 (15.6%) |
| 18-50y | 2354 (54.5%) | 21 (12.1%) | 2333 (56.3%) |
| 50-60y | 609 (14.1%) | 29 (16.8%) | 580 (14%) |
| 60-70y | 376 (8.7%) | 35 (20.2%) | 341 (8.2%) |
| 70-80y | 232 (5.4%) | 42 (24.3%) | 190 (4.6%) |
| 80y≤ | 135 (3.1%) | 46 (26.6%) | 89 (2.1%) |
| Female | 1939 (44.5%) | 50 (28.9%) | 1889 (45.2%) |
| **Comorbidities** |  |  |  |
| *Chronic respiratory diseases* | 481 (11%) | 39 (22.5%) | 442 (10.6%) |
| Chronic obstructive pulmonary disease | 310 (7.1%) | 24 (13.9%) | 286 (6.8%) |
| Other chronic pulmonary disease | 153 (3.5%) | 10 (5.8%) | 143 (3.4%) |
| Pleural effusion | 41 (0.9%) | 4 (2.3%) | 37 (0.9%) |
| *Cardiovascular diseases* | 310 (7.1%) | 57 (32.9%) | 253 (6.1%) |
| Ischemic heart disease | 132 (3%) | 27 (15.6%) | 105 (2.5%) |
| Congestive heart failure | 30 (0.7%) | 11 (6.4%) | 19 (0.5%) |
| Cerebrovascular disease | 57 (1.3%) | 15 (8.7%) | 42 (1%) |
| Peripheral vascular disease | 23 (0.5%) | 7 (4%) | 16 (0.4%) |
| Other cardiovascular diseases | 199 (4.6%) | 41 (23.7%) | 158 (3.8%) |
| *Hypertension* | 627 (14.4%) | 102 (59%) | 525 (12.6%) |
| *Immunosuppression* | 164 (3.8%) | 31 (17.9%) | 133 (3.2%) |
| *Cancer* | 205 (4.7%) | 33 (19.1%) | 172 (4.1%) |
| *Deficiency anemia* | 423 (9.7%) | 18 (10.4%) | 405 (9.7%) |
| *Liver and kidney diseases* |  |  |  |
| Liver disease | 404 (9.3%) | 28 (16.2%) | 376 (9%) |
| Chronic kidney disease | 384 (8.8%) | 86 (49.7%) | 298 (7.1%) |
| End stage renal disease | 85 (2%) | 26 (15%) | 59 (1.4%) |
| *Fluid and electrolyte disorders* | 394 (9.1%) | 37 (21.4%) | 357 (8.5%) |
| *Metabolic diseases* |  |  |  |
| Diabetes | 362 (8.3%) | 58 (33.5%) | 304 (7.3%) |
| Obesity (BMI≥30) | 874 (20.1%) | 73 (42.2%) | 801 (19.2%) |
| *Neurological and cognitive disorders* |  |  |  |
| Neurological disorders | 294 (6.8%) | 57 (32.9%) | 237 (5.7%) |
| Paralysis | 53 (1.2%) | 12 (6.9%) | 41 (1%) |
| Depression | 578 (13.3%) | 53 (30.6%) | 525 (12.6%) |
| Cognitive impairment | 87 (2%) | 28 (16.2%) | 59 (1.4%) |
| *Other* |  |  |  |
| Hospitalization | 931 (21.4%) | 92 (53.2%) | 839 (20.1%) |
| Smoking | 643 (14.8%) | 41 (23.7%) | 602 (14.4%) |
| Current smoker | 514 (11.8%) | 30 (17.3%) | 484 (11.6%) |
| Past smoker | 129 (3%) | 11 (6.4%) | 118 (2.8%) |
| Nursing home | 67 (1.5%) | 23 (13.3%) | 44 (1.1%) |
| Home care | 44 (1%) | 17 (9.8%) | 27 (0.6%) |

**Figure 1.**
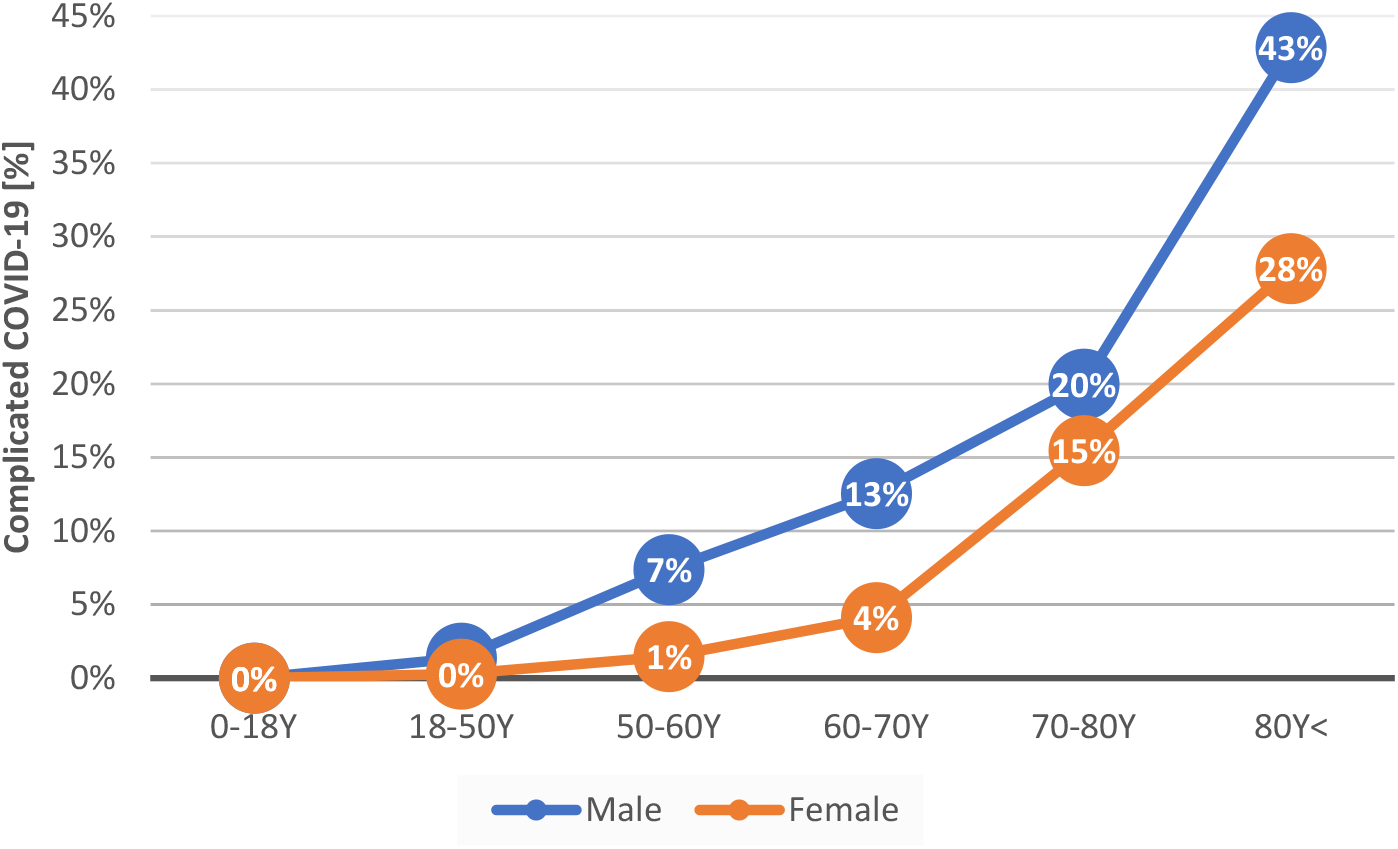
*Age and complicated COVID-19*. Prevalence of complicated COVID-19 (moderate or severe condition, y-axes) in different age groups (x-axis), shown separately for male (blue) and female (orange).

**Table 2.** Association of male sex and COVID-19 complications across age groups.

| Age group | Patient counts<sup>a</sup> | OR<sup>b</sup> | P-value<sup>c</sup> |
| --- | --- | --- | --- |
| 18-50y | (18/1300,3/1033) | 4.77 [1.39, 25.32] | 0.013 |
| 50-60y | (25/314,4/266) | 5.28 [1.79, 21.15] | 0.003 |
| 60-70y | (29/202,6/139) | 3.32 [1.31, 10.03] | 0.013 |
| 70-80y | (27/108,15/82) | 1.36 [0.65, 2.95] | 0.473 |
| 80y≤ | (24/32,22/57) | 1.93 [0.89, 4.26] | 0.145 |

<sup>a</sup> Number of males in the complicated/non-complicated COVID-19 cohorts, followed by the corresponding numbers in females
<sup>b</sup> Odds ratios (ORs) and 95% confidence intervals (in brackets). ORs greater than 1 suggest an increased risk for COVID-19 complications in males
<sup>c</sup> P-values adjusted for multiple testing using Benjamini and Hochberg method [19]. Rows are sorted ascendingly by P-value.

Comparing the prevalence of existing conditions in three age groups between the complicated and non-complicated COVID-19 cohorts, reveals multiple risk factors, including obesity for patient 18-50 years (OR: 11.09, 95% confidence intervals, CI: [4.15, 32.67]; P-value < 10^-4^), Chronic kidney disease for patients 50-65 years (4.06 [1.89, 8.38]; P-value = 0.005); and neurological disorders (2.65 [1.69, 4.17]; P-value < 0.001) for patients 65 years or older (for a complete list, see Table 3 and Supplementary File 1).

**Table 3.** Most statistically significant conditions associated with increased risk of COVID-19 complications in age stratified patient groups.

| Condition | Age group | Patient counts<sup>a</sup> | OR<sup>b</sup> | P-value<sup>b</sup> |
| --- | --- | --- | --- | --- |
| Obesity | 18-50y | (14/356,7/1977) | 11.09 [4.15, 32.67] | 2.36E-05 |
| Depression | 18-50y | (7/229,14/2104) | 4.59 [1.55, 12.3] | 0.032 |
| Hypertension | 18-50y | (4/72,17/2261) | 7.37 [1.76, 23.41] | 0.035 |
| Liver disease | 18-50y | (5/125,16/2208) | 5.51 [1.55, 16.07] | 0.037 |
| Chronic kidney disease | 50-65y | (14/87,27/683) | 4.06 [1.89, 8.38] | 0.005 |
| End stage renal disease | 50-65y | (5/8,36/762) | 13.11 [3.21, 48.19] | 0.006 |
| Neurological disorders | 65y≤ | (54/113,57/317) | 2.65 [1.69, 4.17] | 6.24E-04 |
| Chronic kidney disease | 65y≤ | (70/174,41/256) | 2.51 [1.6, 3.97] | 0.001 |
| Other cardiovascular diseases | 65y≤ | (36/70,75/360) | 2.46 [1.49, 4.05] | 0.006 |
| Cognitive impairment | 65y≤ | (28/52,83/378) | 2.45 [1.4, 4.22] | 0.015 |
| Home care | 65y≤ | (16/22,95/408) | 3.12 [1.47, 6.48] | 0.022 |
| Hypertension | 65y≤ | (82/249,29/181) | 2.05 [1.27, 3.4] | 0.027 |
| Cardiovascular diseases | 65y≤ | (50/129,61/301) | 1.91 [1.22, 2.99] | 0.032 |
| Nursing home | 65y≤ | (20/35,91/395) | 2.48 [1.29, 4.65] | 0.036 |

<sup>a</sup> Number of patients with the condition (cases) in the complicated/non-complicated COVID-19 cohorts, followed by the corresponding numbers in patients without the studied condition (controls)
<sup>b</sup> Odds ratios (ORs) and P-values as defined in Table 2. ORs greater than 1 suggest an increased risk for COVID-19 complications in patients with the noted condition

Stratifying over age (below and above 65 years) and sex (Table 4 and Supplementary File 1), kidney diseases appear as a risk factor in all strata (e.g., OR 3.45 [1.57, 8.06]; P-value = 0.015 in women 65 years or more). Additional risk factors include hypertension in males under 65 years (4.56 [2.35, 8.55]; P-value < 0.001); neurological disorders in females 65 years or older (3.55 [1.68, 7.74]; P-value = 0.008); cognitive impairment (4.18 [1.81, 9.72]; P-value = 0.009) and depression (2.94 [1.55, 5.58]; P-value = 0.014) in males 65 years or older.

**Table 4.**
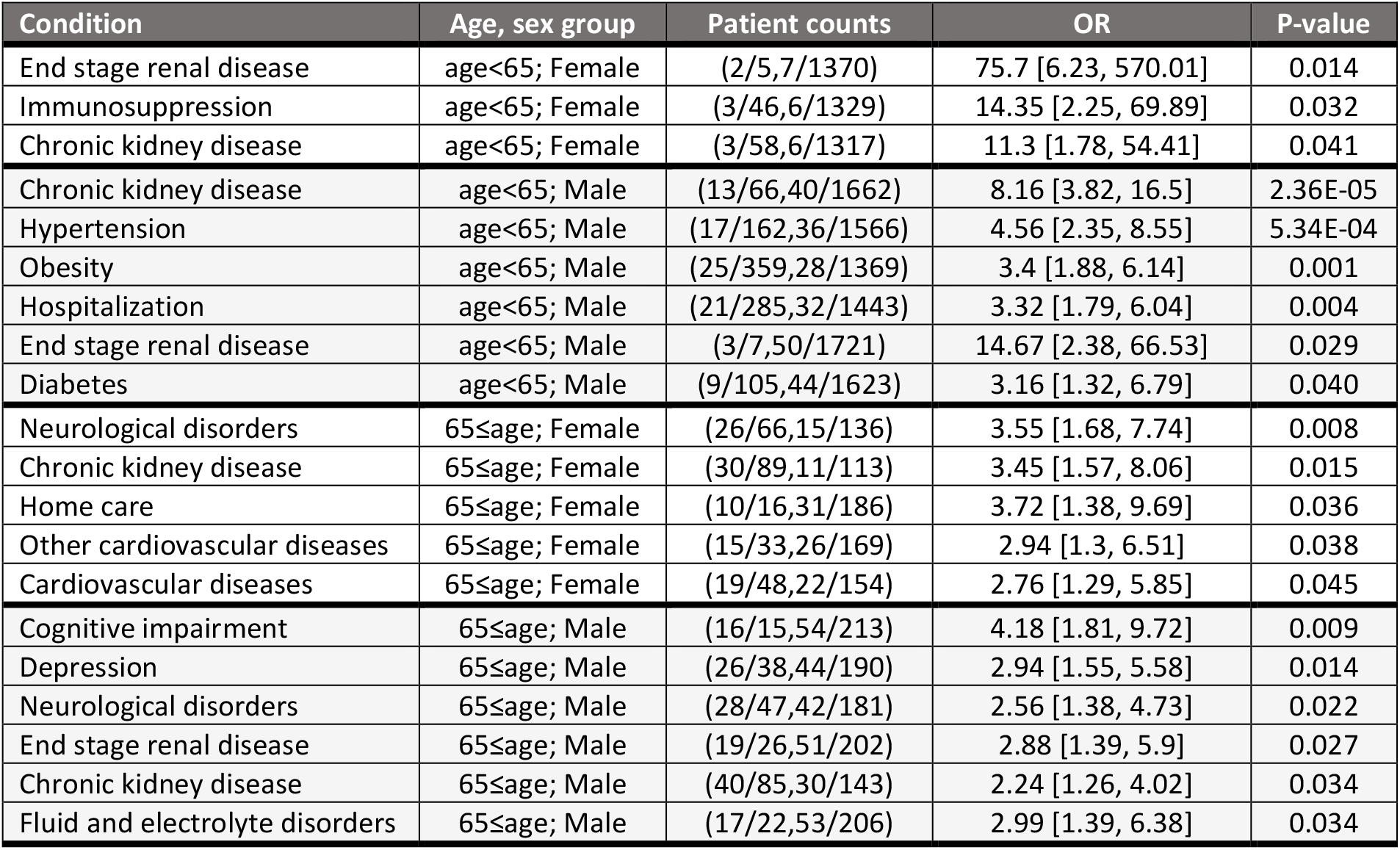
Most statistically significant conditions associated with increased risk of COVID-19 complications in age and sex stratified patient groups. See Table 3 for column information.

| Condition | Age, sex group | Patient counts | OR | P-value |
| --- | --- | --- | --- | --- |
| End stage renal disease | age<65; Female | (2/5,7/1370) | 75.7 [6.23, 570.01] | 0.014 |
| Immunosuppression | age<65; Female | (3/46,6/1329) | 14.35 [2.25, 69.89] | 0.032 |
| Chronic kidney disease | age<65; Female | (3/58,6/1317) | 11.3 [1.78, 54.41] | 0.041 |
| Chronic kidney disease | age<65; Male | (13/66,40/1662) | 8.16 [3.82, 16.5] | 2.36E-05 |
| Hypertension | age<65; Male | (17/162,36/1566) | 4.56 [2.35, 8.55] | 5.34E-04 |
| Obesity | age<65; Male | (25/359,28/1369) | 3.4 [1.88, 6.14] | 0.001 |
| Hospitalization | age<65; Male | (21/285,32/1443) | 3.32 [1.79, 6.04] | 0.004 |
| End stage renal disease | age<65; Male | (3/7,50/1721) | 14.67 [2.38, 66.53] | 0.029 |
| Diabetes | age<65; Male | (9/105,44/1623) | 3.16 [1.32, 6.79] | 0.040 |
| Neurological disorders | 65≤age; Female | (26/66,15/136) | 3.55 [1.68, 7.74] | 0.008 |
| Chronic kidney disease | 65≤age; Female | (30/89,11/113) | 3.45 [1.57, 8.06] | 0.015 |
| Home care | 65≤age; Female | (10/16,31/186) | 3.72 [1.38, 9.69] | 0.036 |
| Other cardiovascular diseases | 65≤age; Female | (15/33,26/169) | 2.94 [1.3, 6.51] | 0.038 |
| Cardiovascular diseases | 65≤age; Female | (19/48,22/154) | 2.76 [1.29, 5.85] | 0.045 |
| Cognitive impairment | 65≤age; Male | (16/15,54/213) | 4.18 [1.81, 9.72] | 0.009 |
| Depression | 65≤age; Male | (26/38,44/190) | 2.94 [1.55, 5.58] | 0.014 |
| Neurological disorders | 65≤age; Male | (28/47,42/181) | 2.56 [1.38, 4.73] | 0.022 |
| End stage renal disease | 65≤age; Male | (19/26,51/202) | 2.88 [1.39, 5.9] | 0.027 |
| Chronic kidney disease | 65≤age; Male | (40/85,30/143) | 2.24 [1.26, 4.02] | 0.034 |
| Fluid and electrolyte disorders | 65≤age; Male | (17/22,53/206) | 2.99 [1.39, 6.38] | 0.034 |

Intriguingly, respiratory diseases and smoking, which contribute to common at-risk definitions (including those by the CDC [1] and the British NHS [3]), while typically more prevalent in complicated COVID-19 patients, are not identified as significant risk factors (e.g., chronic obstructive pulmonary disease in patients 65 years or more: OR 1.36 [0.75, 2.4]; P-value = 0.634); and see Supplementary File 1.

## Discussion

We compared the prevalence of dozens of existing conditions in Israeli SARS-CoV-2 positive and complicated COVID-19 patient cohorts, to highlight those conditions associated with high risk of complications. A few other studies have employed a similar study design to identify risk factors for COVID-19 complications. For example, Ebinger et al [8] studied a cohort of symptomatic COVID-19 individuals (N=442) and examined the association of existing conditions with disease severity; and the OpenSAFELY Collaborative explored the risk of COVID-19-related hospital death among the general population (N>17M). We emphasize that cohort composition dictates the research question it can address: our analysis focuses on SARS-CoV-2 positive individuals, hence searches for risk factors of complications in patients who already contracted the virus (but are potentially asymptomatic), while studying the general population may combine risk factors for infection and COVID-19 severe outcome. Additionally, cohorts that consider only a subset of patients, defined based on disease outcome (e.g., symptomatic or hospitalized) or otherwise non-representative of the entire population (e.g., demographically skewed), may introduce biases to the analysis; instead, we study here all SARS-CoV-2 infected patients in a large, nationwide health organization.

Many conditions highlighted by our analysis have been previously reported [5,6,8] and are part of commonly used at-risk definitions [1,3], including hypertension, obesity, kidney and cardiovascular diseases. We do, however, identify a few additional risk factors, notably depression in patients 18-50 years old and males 65 years or more; and cognitive and neurological disorders in patients 65 years or older. These additions may be, in part, associated with different age distribution in the 65+ years group (median 76y, IQR [70-83.5y] versus 72y [68-78y] in the complicated and non-complicated COVID-19 cohorts, respectively) and rely on small sample size (only seven 18-50y patients with depression in the complicated COVID-19 cohort; Table 3); nonetheless, with some preliminary support [7], they may deserve more consideration in future studies. Our analysis also points out to reduced importance of respiratory diseases and smoking. Both conditions appear as factors in most at-risk definitions [3,5]: Chronic obstructive pulmonary disease has been associated with severe COVID-19 in multiple (though not all [6]) studies [20], while the role of smoking has been somewhat controversial [20,21]. The discrepancies between our analysis and previous reports likely stem from the different cohorts analyzed: SARS-CoV-2 positive individuals, ranging from asymptomatic to severe COVID-19 versus hospitalized COVID-19 patients, respectively. Other study-related attributes, for example country-specific characteristics, may also contribute to the varying significance of the studied risk factors.

In parallel to the COVID-19 epidemiological characterization efforts, researchers have also attempted to use retrospective observational data to derive risk models for severe COVID-19 patients [22]. Such models require ample data of COVID-19 patients for both model training and performance assessment. As, currently, such data are scarce, some models compromised on using data for other diseases with, supposedly, similar clinical trajectory and complications. For example, DeCapprio et al [19] trained models on US Medicare claims data to predict inpatient visits with a primary diagnosis of either pneumonia, influenza, acute bronchitis, or other specified upper respiratory infections as proxy for COVID-19 complications. However, as previously reported (e.g., in [24]), and in agreement with our analysis, severe COVID-19 patient characteristics differ considerably from other diseases’, thus limiting the generalizability of such models to COVID-19 and requiring adjustments of their parameters [4].

Our study has several limitations. First and foremost, the number of complicated COVID-19 patients in MHS data is below 200, limiting the statistical power of our analysis. Second, healthcare policies and, in particular, testing criteria, may systematically bias the composition of SARS-CoV-2 positive cohort. Third, asymptomatic and mild COVID-19 patients (currently in the non-complicated cohort) may deteriorate and eventually be part of the complicated cohort, potentially modifying the results of the analysis. Fourth, our analysis is univariate in nature, testing the association of individual conditions with COVID-19 complications; as such, it is unable to uncover more complex relations, e.g., interdependencies between conditions and COVID-19 complications. Finally, we focused on data from Israel; characteristics in other geographies may differ [24]. We attempted to mitigate some of these limitations by age and sex stratification and robust estimations of statistical significance. We also note that, at the current point in time, many of these shortcomings are shared by all published COVID-19 research work.

Notwithstanding these limitations, our work adopts a novel vantage point to the problem of identifying patients at increased risk for COVID-19 complications. Importantly, as SARS-CoV-2 containment efforts focus on patients at risk for severe complications (for example, shielding vulnerable population in the UK [3]), changes in the list of considered conditions may have huge effect on a large number of individuals, thus calling for continuous fine-tuning of the corresponding definitions.

## Supplementary Information

*Supplementary File 1. Odds ratio analysis*.

## Ethical approval

The study was approved by Maccabi Health Services’ institutional review board.

## Data Availability

Patient-level de-identified data from Maccabi Health Services

## Acknowledgement

We thank Guy Amit and Irena Girshovitz, KI Research Institute, for insightful discussions and comments.

## Author Contributions

**Conceptualization: NK; Data Curation: BM, KM, YB; Investigation and methodology: CY, BM, NK; Project Administration: PA, VS; Writing – original draft preparation: CY; Writing – review and editing: CY, BM, KM, PA, YB, GC**.

